# Shared residual risk of childhood undernutrition indicators in Malawi. An application of shared spatial component model

**DOI:** 10.1101/2020.09.19.20197772

**Authors:** Alfred Ngwira

## Abstract

Many studies have looked at the residual risk of the specific child undenutrition indicators. This study aimed at mapping the shared risk of two of the undernutrition indicators. The shared spatial component model was fitted to two of the child undernutrition indicators using 5066 child records of the 2015 Malawi demographic health survey data. The spatial components were modelled by the convolution prior, with the structured components being assigned the conditional autoregressive distribution. The southern region is at the greatest risk of having stunting and wasting, wasting and underweight, as compared to the central and northern region. The shared risk of stunting and underweight is randomly distributed. Interventions to reduce the shared risk of child undernutrition should focus on the southern region and a little bit in the central region, and attention should be on addressing the issue of overpopulation and effects of climate change.

## Introduction

Child undernutrition is a form of malnutrition resulting from eating less. Some forms of undenutrition are stunting, wasting and underweight. Child undernutrition is said to be associated with poor survival, poor physical and cognitive development (UNICEF, 2013) and obesity later in life (WHO, 2017). The global burden of childhood malnutrition as of 2018 (UNICEF, WHO and World Bank, 2019) was 21.9% for stunting, 7.3% for wasting and 5.9 % for overweight. In Malawi, it is estimated that 37% of under five children are stunted, 3% are wasted, 12% are underweight and 4.5 % are overweight, according to the 2015 Malawi demographic health survey (MDHS) report (NSO, 2017).

The child undernutrition status is measured by the anthropometric measures (WHO, 2006). Stunting is measured by height-for-age z score (HAZ) and is a sign of chronic food inadequacy. Wasting is measured by weight-for-height z score (WHZ) and is a manifestation of acute situation related to illness or lack of food. Underweight is measured by weight-for- age z score (WAZ) and it is a result of either wasting or stunting or both. The three undernutrition indicators are likely to be correlated considering that they share the same biomarkers of the same child, for example height and weight. Correlation studies have confirmed pairwise correlation between the undernutrition indicators particularly between underweight and stunting, and between underweight and wasting when using cross sectional data (Ngwira et al, 2017; Kassie and Workie, 2019). Wasting has been observed to be associated with stunting when the data is from a longitudinal study (Richard et al, 2012). Most studies on the distribution of the residual risk of the child undernutrition status have focused on the risk of one indicator without looking at the common risk shared by the indicators (e.g Kandala et al, 2009; Habyarimana et al, 2016;.Alemu et al, 2016; Khan and Mohanty, 2018; Bharti et al, 2019; Hagos et al, 2017). In addition, most of these studies have used univariate modeling, forgetting that undernutrition indicators are correlated which requires the use of the joint modeling. One problem with the univariate model of correlated outcomes like the undernutrition indicators is the over estimation bias attached to parameter estimates which can result in wrong statistical inference (Fang et al, 2016) and hence forth wrong policy making. One study on shared spatial heterogeneity regarding child undernutrition status has been spotted though (Kinyoki et al, 2016), and others have also been seen on other health outcomes (Manda, 2011; Kazembe et al, 2009; Kandala et al, 2014) and none on child undernutrition status has been seen for Malawi to my knowledge.

The aim of this study was to investigate co-distribution of the residual risk of the child undernutrition status indicators so as to determine the areas where the indicators are strongly correlated and to hypothesize possible common risk factors for further epidemiological investigation.

## Materials and Methods

### Data

The study used the 2015 MDHS data which was downloaded from the DHS website after being given permission. The focus was on children who were less than five years of age. Since the study used the secondary data, the Malawi Health Research Committee saw it inappropriate to grant the ethical approval. The MDHS study according to NSO (2017) was a two stage cluster sampling with stratification where clusters were stratified by residence (urban/rural) and then in each cluster, households were randomly selected. In the first stage, 850 clusters, comprising of 173 clusters in urban areas and 677 clusters in rural areas were selected by probability proportional to size (PPS) cluster sampling method. In the second stage, 30 households from each urban cluster and 33 households from each rural cluster were selected by systematic sampling. The data from households was then collected using four questionnaires that is, the woman, man, household and then biomarker questionnaire. The response variables of interest considered were child undernutrition status indicators: stunting (HAZ<-2/HAZ≥-2), wasting (WHZ<-2/WHZ≥-2), and underweight (WAZ<-2/WAZ≥-2), and the independent variables were child age (in months), mother body mass index (kg/m^2^), child sex (male/female), education (no education/primary/secondary), wealth quantile (poorest/poor/rich/richer/richest) and district of the child.

### Statistical analysis

Bivariate scatter plots were made between two of the three undernutrition indicators to asses correlation. A multiple variable joint shared heterogeneity model of any of the two undernutrition status indicators was then fitted. Specifically, a joint shared spatial model of stunting and wasting, stunting and underweight, and wasting and underweight were fitted. Theoretically, a bivariate shared spatial model of the two Bernoulli distributed health outcomes is defined as follows according to Manda et al (2012). Let *π*_*ij*1_ be the probability of child *i* in area *j* of having disease of the first kind, and *π*_*ij*2_ the probability of child *i* in area *j* of having disease of the second kind. Then the joint shared spatial model of the two diseases is defined as, log *it*(*π*_*ij*1_) = *α*_1_ + *X*^*T*^*β*_1_ + *ϕ*_*j*_*κ* + *φ*_*j*1_ and log *it* 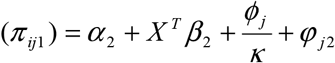 where *X* is a vector of fixed effects and *ϕ*_*j*_ is the area level shared spatial component and *φ*_*j*1_ and *φ*_*j*2_ are the two area level spatial effects which are disease specific. The parameter *κ* represents the differential gradient of the shared spatial component of the two diseases. The ratio of the scaling parameters, *κ* to 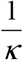 compares the *β* weight of disease 1 relative to disease 2 associated with the shared component. The shared spatial effect, *ϕ*_*j*_ represents the proxy of the area level unmeasured risk factors influencing both diseases and the disease specific spatial effects, *φ*_*j*1_ and *φ*_*j*2_ denote the proxies for the unmeasured risk factors specific to the two diseases. The focus of this study was on the investigation of the shared spatial component which represents the shared residual risk between the two diseases. In this case, the shared spatial component was the district of the child, where the shared risk would be observed per district. The rest of the independent variables were used as control variables. Model estimation was fully Bayesian using the Gibbs sampling where model parameters were assigned prior distributions. All fixed effect parameters were assigned the normal distribution with a large variance assuming prior ignorance. All the spatial components were assigned the convolution prior where they were split into structured and unstructured, that is, *u* + *v*. The unstructured spatial components were assigned the normal distribution with zero mean, that is, 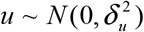 and the structured spatial effects were modeled by the intrinsic conditional autoregressive (ICAR) normal distribution (Besag et al, 1991), that is, 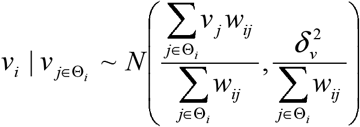 where *w*_*ij*_ was the weight relating adjacent areas, that is, if *w*_*ij*_ = 1 then the adjacent areas *i* and *j* were neighbors and if *w*_*ij*_ = 0, then the adjacent areas were not neighbors. The idea behind the CAR prior for the area spatial effect is that the spatial effect of the area is the average of spatial effects of the neighbors of the given area. The variance parameters were assigned the gamma distribution. The models were fitted by WinBUGS using R2WinBUGS package in R. A total of 25000 iterations were used and after a burn in of 10000 iterations and thinning of every 30^th^ iteration, 500 iterations were left for posterior analysis. Before posterior analysis, the Markov chain Monte Carlo (MCMC) chains for all parameters were assessed for convergence to the posterior distribution by CODA package in R. The Geweke, Gelman-Rubin, and Heidelberger-Welch diagnostic tests confirmed the convergence of MCMC chains. Posterior analysis involved the mapping of the posterior mean of the shared spatial component and the posterior probability that the shared spatial component for each district was more than one.

## Results

Figure 1 presents bivariate correlation of the three malnutrition indicators z scores, that is, the height-for-age, weight-for-age, and weight-for-height. There is linear correlation between weight-for-age and height-for-age z score, and between weight-for-height and weight-for-age z score since the scatter plots form a linear pattern. There is a very weak correlation between weight-for-height and height-for-age z score since the pattern in a scatter plot cannot be clearly defined. A shared spatial model of stunting and wasting, stunting and underweight, and then wasting and underweight was then fitted. Table 2 shows the estimates of the variance parameters.

**Figure 1:**
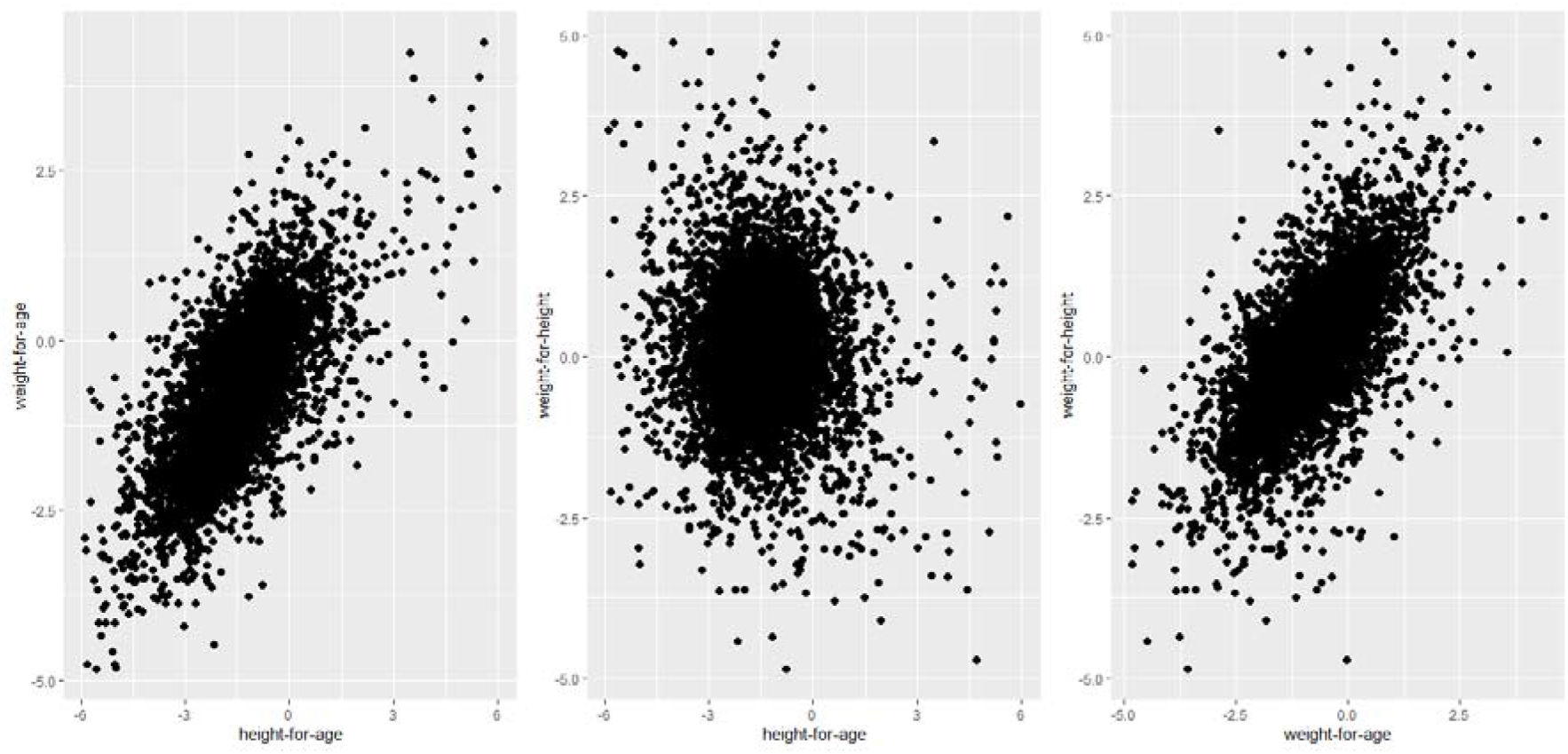
Bivariate correlation of the z scores for the malnutrition indicators

The proportion of variance due to shared risk factors between the undernutrition indicators is very high for wasting and underweight (84% and 71%) and is moderate for stunting and wasting (44 % and 19%), and stunting and underweight (46 % and 40 %). The scaled shared spatial variance parameters for wasting and underweight (0.174 and 0.033) are relatively higher than those of stunting and wasting (0.007 and 0.018) and stunting and underweight (0.008 and 0.015) which are close to zero. This is suggesting insignificant shared spatial variation regarding the latter two pairs of undernutrition status indicators. Figure 2 shows the shared spatial pattern of the two of the three undernutrition status indicators. The shared risk pattern of stunting and wasting (Figure 2, a) shows many areas in the southern region being at increased risk to both stunting and wasting. One region in the north west, called Mzimba is also at increased risk to both stunting and wasting. The shared unobserved risk pattern of stunting and underweight (Figure 2, b), shows the high risk areas being randomly distributed across the country. Similar to the shared risk pattern of stunting and wasting, the shared risk pattern of wasting and underweight (Figure 2, c) shows high risk areas being clustered in the south. The posterior probability map regarding stunting and wasting (Figure 2, d) that the estimated risk ratio is greater than one is showing many areas in the center and south having high probability that the risk ratio exceeds one. With regard to stunting and underweight (Figure 2, e), many areas across Malawi, regardless of region, have high probability that the risk ratio is greater than one. Regarding wasting and underweight (Figure 2, f), most areas in the southern region have a higher probability that the shared risk ratio is more than one. Spatial cluster analysis by the Moran I statistic, shows that there is significant clustering regarding shared risk to stunting and wasting (Moran I statistic=0.464, p-value = 0.009). There is also significant spatial autocorrelation regarding unobserved common risk to wasting and underweight (Moran I=0.392, p-value = 0.026). The unobserved shared risk to both stunting and underweight is randomly distributed across Malawi (Moran I statistic=- 0.044, p-value = 0.539).

**Table 1:**
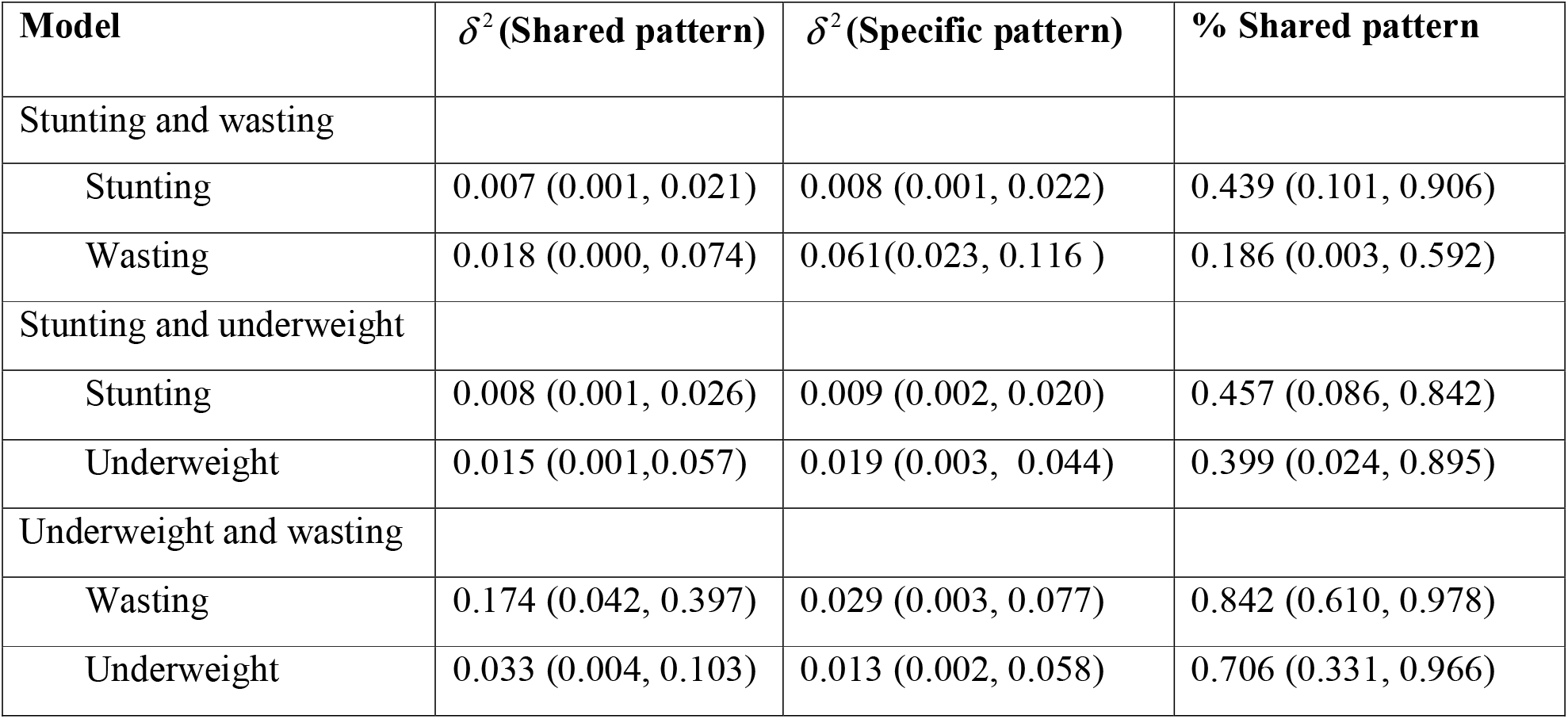
Variance parameters of the shared spatial component model

| Model | $\delta^2$ (Shared pattern) | $\delta^2$ (Specific pattern) | % Shared pattern |
| --- | --- | --- | --- |
| Stunting and wasting |  |  |  |
| Stunting | 0.007 (0.001, 0.021) | 0.008 (0.001, 0.022) | 0.439 (0.101, 0.906) |
| Wasting | 0.018 (0.000, 0.074) | 0.061(0.023, 0.116 ) | 0.186 (0.003, 0.592) |
| Stunting and underweight |  |  |  |
| Stunting | 0.008 (0.001, 0.026) | 0.009 (0.002, 0.020) | 0.457 (0.086, 0.842) |
| Underweight | 0.015 (0.001,0.057) | 0.019 (0.003, 0.044) | 0.399 (0.024, 0.895) |
| Underweight and wasting |  |  |  |
| Wasting | 0.174 (0.042, 0.397) | 0.029 (0.003, 0.077) | 0.842 (0.610, 0.978) |
| Underweight | 0.033 (0.004, 0.103) | 0.013 (0.002, 0.058) | 0.706 (0.331, 0.966) |

**Figure 2:**
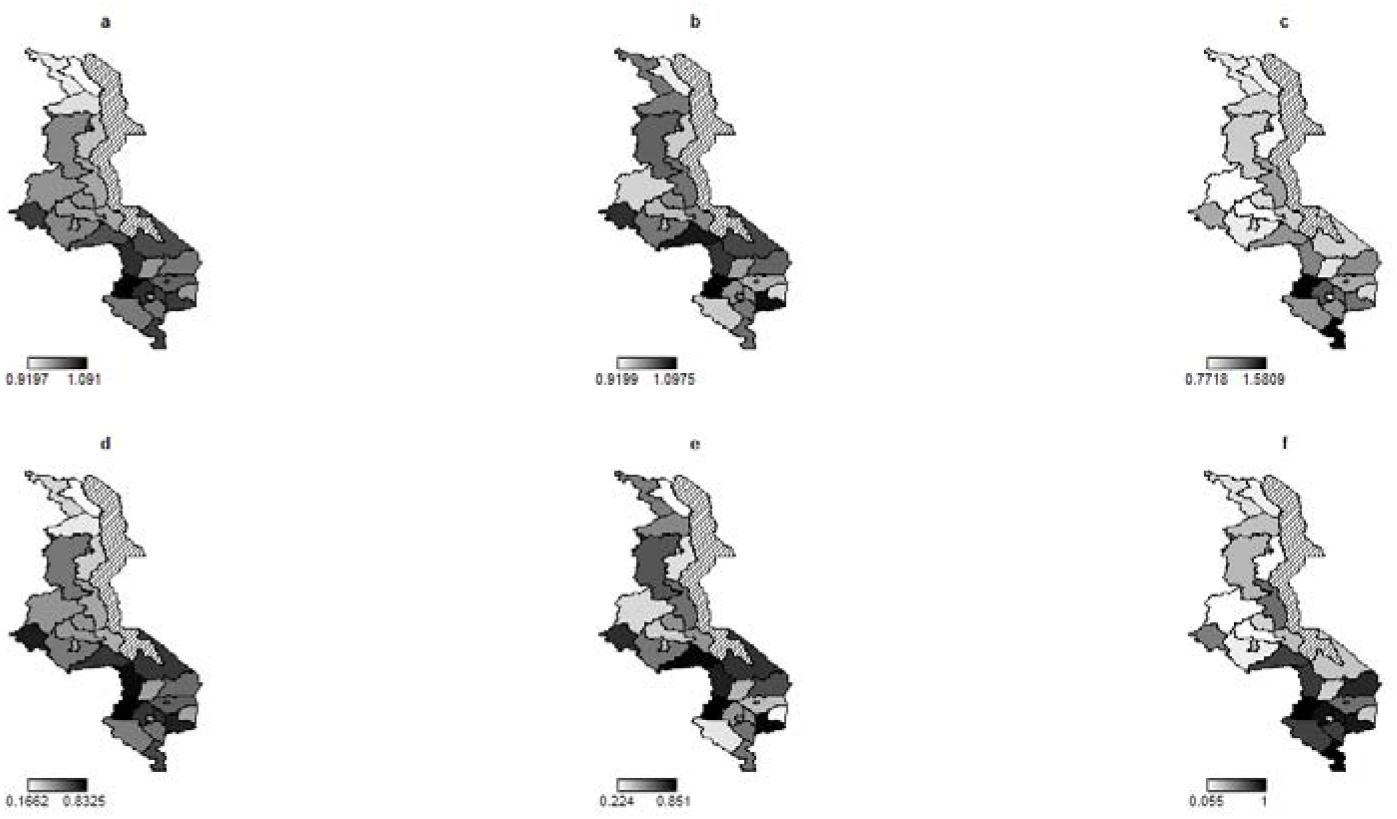
The shared residual risk of undernutrition (a – c). (a) stunting and wasting, (b) stunting and underweight, (c) underweight and wasting. Probability that RR > 1 (d – f). (d) stunting and wasting, (e) stunting and underweight, and (f) underweight and wasting.

## Discussion

The study aimed at exploring the bivariate shared residual risk pattern of the child undernutrition indicators using the shared spatial component model. The strength of the study is in the use of the nationally representative data which would permit inferences about the whole nation. In addition, the analytical procedure is relatively novel as the routine method tends to use the separate models to investigate the residual risk of one undernutrition indicator.

The study finds that most areas in the southern region including few in the central region are consistently being at increased risk to all the three pairs of undernutrition indicators. The findings of the study are consistent with the literature (Nube and Sonneveld, 2005; Kinyoki et al, 2020) where it was found that the distribution of underweight and stunting separately were more prevalent in the south and center than in the northern region. One possible explanation to this spatial gradient would be the effect of population density. The southern region has the highest population density (244 people/km^2^) seconded by the central region (211 people/km^2^) and lastly the northern region with low density (84 people/km^2^) (NSO, 2018). The observed spatial gradient of the shared risk to undernutrition indicators, where the high risk coincides with the high population density is supported by the Nube and Sonneveld (2005) where they too found that underweight hot spots in Africa were also high population density areas. The population density though itself is not correlated with undernutrition, but the pressure on land and its deterioration in quality due to high population brings about poor nutrition conditions (Sonneveld et al, 2003).

The other possible driver of the observed shared spatial structure would be the climate. The main factors of climate are rainfall and temperature. High temperature tends to be associated with increased risk of undernutrition (Kinyoki et al, 2016). The effect of temperature on malnutrition is due to the fact that temperature is directly linked to aridity according to Quan et al (2013), which in turn has an impact on malnutrition (Grace et al, 2012). On the other hand, high rainfall has been found to be associated with increased risk of stunting, and low rainfall is associated with increased risk of wasting and underweight (Ngwira, 2020; Kinyoki et al, 2016). The high risk to undernutrition indicators manifested in the southern region in Malawi is therefore as expected, as the south eastern part (Zomba, Mulanje and Thyolo) is associated with high rainfall and the south western part including the southern tip (Balaka, Chikwawa and Nsanje) is associated with high temperature and low rainfall (Vincent et al, 2014). The effect of climate change like flooding would also be the other contributing factor to the observed high risk in the southern region. Areas especially in the southern tip experience flooding from Shire river almost every year. Flooding has been documented to enhance food insecurity by reducing fish catch rates due to dilution of fish in greater volumes of water (Tregidgo et al, 2020). Flooding is also associated with child morbidity like diarrhoea which in turn is associated with high risk to undernutrition (Poda et al, 2017).

The weakness of this study though is that the data is a little bit out dated and hence the residual risk patterns might have changed over the last five years. Nonetheless, the observed shared risk patterns may guide the policy makers regarding the areas with increased shared risk in the absence of up to date nationally representative data, as currently there is no up to date data to my knowledge.

## Conclusion

The study finds non random pattern of the shared risk to stunting and wasting, wasting and underweight, where the southern region is at increased risk compared to the central and northern region. There is no significant clustering regarding the shared risk to stunting and underweight. The observed south to north gradient of spatial risk pattern calls for epidemiologist to further investigate the actual shared risk factors bringing about this spatial risk gradient. In addition, nutrition intervention policies should include interventions to address the effects of climate and overpopulation on undernutrition status.

## Data Availability

Data is available upon request from the corresponding author.

## Contributions

AN, conceptualized the study, prepared and analysed the data, drafted and reviewed the manuscript.

## Consent for publication

This study used secondary data, the 2015 MDHS data which was downloaded from the website (www.dhsprogram.com/dataset_admin) after being granted permission. No separate permission was required for usage and publication.

## Acknowledgements

The author acknowledges the demographic health survey (DHS) for providing the data for analysis.

